# The acceptability, appropriateness, and equity of remote consultations in sexual and reproductive health services in England and Wales: a qualitative study

**DOI:** 10.64898/2026.02.06.26345738

**Authors:** Tom Witney, Charlotte Spurway, Jo Gibbs, Helen Munro, Iestyn Williams, Danielle Solomon, Melvina Woode Owusu, Jo Josh, Andrew Copas, Jonathan DC Ross, Louise Jackson, Fiona Burns

## Abstract

**Background:** Remote consultations (via telephone or video) were critical during COVID-19 restrictions for Sexual and Reproductive Health Services (SRHS) in England and Wales and continue to be implemented widely. However, there remains limited evidence on their impact on outcomes and health inequalities. This study, guided by the Framework for Digital Health Equity, explores the equity implications of remote sexual health consultations in England and Wales, examining their acceptability and appropriateness.

**Methods:** We conducted semi-structured interviews with 54 service users and potential service users and 35 professional stakeholders across three diverse case study areas. Participants were purposively sampled to ensure diversity in socio-demographic profile and service experience. All participants provided informed consent, and ethical approval for the study was granted by NHS Research Ethics Committee (REC: 23/NS/0128). Data were thematically analysed into narrative themes, with findings interpreted collaboratively with public and patient involvement groups.

**Results:** The study found that remote consultations have reshaped care delivery, improving efficiency but also generating additional clinical workload and, for some clinicians, reduced job satisfaction. While generally acceptable, remote models were perceived to affect rapport, extend care pathways for some service users, and impact privacy and safeguarding. We found the benefits of remote consultation are unevenly distributed, with those experiencing digital exclusion, who lack access to suitable private spaces, or who experience language barriers, being less able to take advantage of their convenience.

**Conclusions:** Remote consultations transform SRHS delivery, improving efficiency for some but introducing challenges by impacting interaction, equity, and confidentiality. Benefits and burdens are unevenly distributed, reflecting structural, social, and individual factors influencing access. SRHS must ensure equitable access to appropriate care for all populations when implementing remote consultations. The findings show that a one-size-fits-all approach is not appropriate, and that giving service users choice in how they consult, including in-person, is important.

## INTRODUCTION

The NHS 10-year plan calls for greater use of digital technology to improve access to care (1). In the context of a decade of funding reductions and rising demand, sexual and reproductive health services (SRHS) have reconfigured delivery models, increasingly relying on digital tools (2). Online pathways for sexually transmitted infection (STI) testing are now routine (3), and the COVID-19 pandemic accelerated the expansion of remote consultation methods in SRHS and other healthcare settings (3–5). These developments are not discrete interventions, but part of broader system change driven to a large extent by fiscal constraint. In some instances, implementation has progressed alongside generation of evidence to guide the process (3), and ongoing assessment and evaluation are therefore essential.

The introduction of remote consultations within healthcare has been reported to increase convenience and efficiency (6, 7). Remote consultations have the potential to benefit people where clinic-based services are not easily accessible, such as in rural communities and people with disabilities or caring responsibilities (6). They have also been reported to improve waiting times and potentially reduce costs (6, 8). However, there are concerns about the increased use of remote consultations in SRHS. These include challenges with confidentiality and privacy, the inability to conduct physical examinations, and concerns about patient acceptability (9). Implementing remote consultations may also decrease patient satisfaction, and issues of digital inequity persist (6, 8, 10). In a study of contraception providers in the US, the introduction of remote consultations changed ways of working and created new demands on services, highlighting the need for appropriate protocols and IT infrastructure to support this shift (11).

The expansion of remote consultations can exacerbate existing inequalities, particularly where these characteristics intersect, such as older adults with limited income and digital literacy (12). A recent systematic review examining the impact of remote consultations on SRHS found that while they enabled continued access during periods of reduced in-person provision, experiences were not equal across all service users (9). For some, remote consultations offered convenience and reduced stigma; for others, they reinforced existing barriers. A key equity concern with remote SRHS is digital exclusion, as unequal access to devices, connectivity, and digital literacy disproportionately affects marginalised populations, including those on low incomes and older adults, those already at greater risk of poor SRH outcomes (8, 13).

This paper is grounded in the Framework for Digital Health Equity, which provides a tailored model to guide equitable digital technology provision (Figure 1) (14). It was developed by extending the existing National Institute on Minority Health and Health Disparities research framework to include digital determinants of health (DDoH) including technology access, digital literacy, and community infrastructure. These determinants operate across societal levels to influence health, functioning, quality of life, and equity in digital healthcare access, outcomes, and design (15). This framework was selected over alternative equity and access models which focus primarily on healthcare utilisation, such the Levesque, Harris & Russell (2013) model of patient-centred access to healthcare (16), or those that are less specific about digital mechanisms, such as the Health Equity Framework (17), and the Socioecological Model of Health (18). Its emphasis on the DDoH embedded in a broader social infrastructure helped us to explore the influence of remote consultations across social contexts and infrastructure levels.

**Figure 1.**
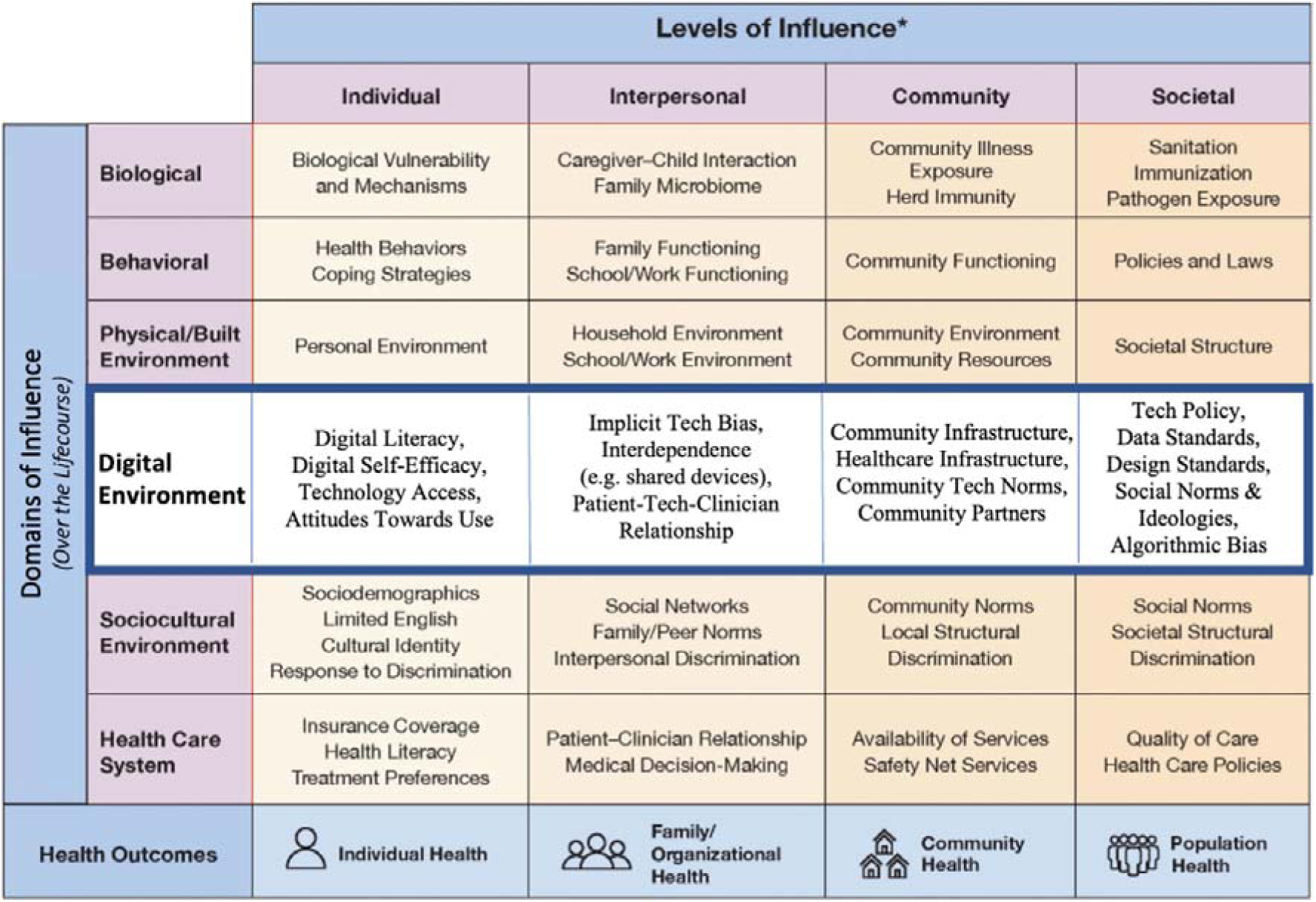
Framework of Digital Health Equity. Figure adapted from: Richardson, S., Lawrence, K., Schoenthaler, A.M. et al. A framework for digital health equity. NPJ Digit. Med. 5, 119 (2022). https://doi.org/10.1038/s41746-022-00663-0. License under CC BY: https://creativecommons.org/licenses/by/4.0/

This study forms part of the CONNECT study (NIHR153151), a wider investigation of the impact of remote consultations in SRHS on health inequalities in relation to access to care, clinical outcomes, timeliness of treatment, the identification of domestic violence/sexual violence and cost-effectiveness. The aim of this qualitative analysis was to explore the equity implications of remote sexual health consultations in England and Wales, examining their acceptability and appropriateness across diverse populations.

## MATERIALS AND METHODS

### Study design

We conducted a qualitative study using semi-structured in-depth interviews with 54 service users (or potential service users) and 35 professional stakeholders across three case study areas (CSAs). The three CSAs were selected for sociodemographic diversity, and to explore differences in rural and urban settings. Case study sites also offered diverse service delivery models (Table 1).

**Table 1.** Overview of appointment and consultation options by case-study area.

|  | **CSA 1** | **CSA 2** | **CSA 3** |
| --- | --- | --- | --- |
| Walk-ins | Under 25s | ☒ | X |
| Booked appointments | ☒ | ☒ | ☒ |
| Online postal self-sampling | X | ☒<br>(in-house provision) | ☒<br>(external provider) |
| Face to face consultations | ☒ | ☒ | ☒ |
| Remote consultations | Limited telephone appointments | ☒ | ☒ |

Within this study, a remote consultation refers to synchronous care conducted between patients and clinicians without in-person contact, and can take the form of telephone, video conferencing, or secure two-way written communication via SMS, email, or online messaging platforms (19). We excluded asynchronous consultations taking place as part of online postal self-sampling (OPSS) as this has been explored elsewhere (3). In this study, SRH services include in-person STI testing, in-person and remote treatment, contraception, advice giving, HIV pre-exposure prophylaxis (PrEP), and abortion care.

### Sampling and recruitment

We employed maximum variation sampling to include a diverse service user sample, including age, disability, educational background, ethnicity, experience with SRHS, gender, sexuality,,, and socioeconomic status, including those who had not engaged with SRHS (20). We purposively sampled professional stakeholders, including doctors, nurses, healthcare assistants, counsellors, managers and commissioners, to include a range of roles from across the CSAs. The target sample comprised 15 to 20 service users/potential services users and approximately 10 to 15 professional stakeholders in each CSA. Participants (both service users and professional stakeholders) were recruited through posters displayed in SRH clinics, direct in-person approaches within clinic settings, outreach via established networks, email invitations, text messages sent to recent service users, social media outreach, and the CONNECT study website.

Service users/potential service users were eligible to participate if they were aged 16 years or older and lived within one of the CSAs. Participants were also required to have the capacity to provide informed consent and to speak and read English. Eligibility criteria for professional stakeholders included being currently involved in the delivery of SRHS within the three CSAs.

### Data collection

To maximise engagement, participants could be interviewed either in-person at one of the SRHS health centres involved in the study, on university premises, over the phone or via Microsoft Teams or Zoom. A screening questionnaire was used to gather socio-demographic data and promote sample diversity. Semi-structured interviews were conducted between March 2024 and January 2025 by CS and TW.

Interviews used a topic guide that focused on experiences with SRHS, including service accessibility, such as booking and attending appointments and remote vs. face-to-face consultations. Those who had not accessed SRHS discussed barriers and any alternative services they had engaged with. Interviews explored participants’ opinions and perceptions of the impact of remote consultations on health inequalities and gathered recommendations for improving access. Service users were given a £20 shopping voucher in appreciation of their time.

For professional stakeholders, interviews involved discussions about perspectives on SRH consultations, both remote and face-to-face. Participants with consultation experience discussed their roles, consultation processes, and the benefits and challenges of remote consultations, including impacts on service delivery, safeguarding, and patient interaction. Those without direct experience shared their perceptions of the impact of remote consultations on their colleagues and on healthcare access. The interviews also examined how remote consultations affect health inequalities and identified training and support needs for both clinicians and service users.

### Data analysis

Audio recordings were transcribed verbatim by a university-approved transcription company and pseudonymised prior to analysis. Data were thematically analysed, with findings interpreted collaboratively with public and patient involvement groups. Data were coded by TW and CS using a code book developed after double-coding a sample of 10 transcripts. During coding, TW and CS met weekly to discuss additional codes that had been generated inductively and add them to the code book. Themes were developed and refined through discussion. NVivo software v.14 (Lumivero, Burlington, MA) was used to support data management and facilitate the analysis process.

Ethical approval for this study was granted by NHS Research Ethics Committee (REC: 23/NS/0128). All participants provided informed consent prior to participation.

## FINDINGS

A total of 54 service users/potential service users participated in this study. The characteristics of these participants are presented in Table 2. Thirty-five interviews were conducted with professional stakeholders, whose characteristics are summarised in Error! Reference source not found..

**Table 2.** Service user self-reported characteristics.

|  |  | CSA 1 | CSA 2 | CSA 3 | Overall |
| --- | --- | --- | --- | --- | --- |
| Age (years) | 16-20 | 2 | 3 | 3 | 8 |
|  | 21-24 | 2 | 4 | 2 | 8 |
|  | 25-34 | 10 | 5 | 8 | 23 |
|  | 35-44 | 1 | 2 | 3 | 6 |
|  | 45+ | 6 | 3 | 0 | 9 |
| Gender | Cis man | 8 | 4 | 4 | 19 |
|  | Cis woman | 9 | 10 | 8 | 23 |
|  | Trans man | 1 | 0 | 1 | 2 |
|  | Trans woman | 2 | 0 | 1 | 3 |
|  | Non-binary | 1 | 2 | 1 | 4 |
|  | Prefer not to say | 0 | 1 | 1 | 2 |
| Ethnicity | Asian or Asian British | 2 | 4 | 0 | 6 |
|  | Black/African/Caribbean/Black British | 3 | 4 | 0 | 7 |
|  | Mixed/multiple ethnic groups/other | 3 | 1 | 2 | 6 |
|  | White | 13 | 8 | 14 | 35 |
|  | Prefer not to say | 0 | 0 | 0 | 0 |
| Sexuality | Heterosexual or straight | 9 | 6 | 9 | 24 |
|  | Gay or Lesbian | 6 | 7 | 2 | 15 |
|  | Bisexual | 6 | 2 | 3 | 11 |
|  | Other | 0 | 2 | 2 | 4 |
|  | Prefer not to say | 0 | 0 | 0 | 0 |
| Disability** | Yes | 7 | 8 | 3 | 18 |
|  | No | 14 | 9 | 12 | 35 |
|  | Prefer not to say | 0 | 0 | 1 | 1 |
| Education | Secondary or earlier | 1 | 0 | 1 | 2 |
|  | Further education | 3 | 4 | 4 | 11 |
|  | University/postgraduate | 17 | 13 | 11 | 41 |
|  | Prefer not to say | 0 | 0 | 0 | 0 |
| Ever used<br>SRHS services | Yes | 20 | 12 | 11 | 43 |
|  | No | 1 | 4 | 5 | 10 |
|  | Prefer not to say | 0 | 1 | 0 | 1 |
| Prior<br>diagnosis of<br>an STI | Yes | 10 | 3 | 2 | 15 |
|  | No | 11 | 14 | 13 | 38 |
|  | Prefer not to say | 0 | 0 | 1 | 1 |
| **Overall** |  | **21** | **17** | **16** | **54** |

\*Participants responded to the question “Do you have any physical or mental health conditions or illnesses lasting or expected to last 12 months or more?”

**Table 3.** Professional stakeholders self-reported characteristics.

|  |  | CSA 1 | CSA 2 | CSA 3 | Overall |
| --- | --- | --- | --- | --- | --- |
| Role | Consultant/Medical Doctor | 4 | 5 | 1 | 10 |
|  | Nurse/Health Advisor | 4 | 3 | 4 | 11 |
|  | Healthcare Assistant | 1 | 0 | 2 | 3 |
|  | Counsellor | 0 | 1 | 0 | 1 |
|  | Manager | 1 | 3 | 1 | 5 |
|  | Commissioner | 1 | 2 | 0 | 3 |
|  | Other | 1 | 0 | 1 | 2 |
| Gender | Man (including trans man) | 7 | 2 | 0 | 9 |
|  | Woman (including trans woman) | 5 | 12 | 8 | 25 |
|  | Non-binary | 0 | 0 | 0 | 0 |
|  | Prefer not to say | 0 | 1 | 1 | 1 |
| Ethnicity | Asian or Asian British | 2 | 3 | 0 | 5 |
|  | Black/African/Caribbean/Black British | 1 | 2 | 0 | 3 |
|  | Mixed/multiple ethnic groups | 0 | 0 | 0 | 0 |
|  | White | 9 | 8 | 9 | 26 |
|  | Other ethnic group | 0 | 0 | 0 | 0 |
|  | Prefer not to say | 0 | 1 | 0 | 1 |
| **Overall** |  | **12** | **14** | **9** | **35** |

Initial findings were developed in dialogue with the overarching themes of appropriateness, acceptability, and equity. These initial themes and subthemes were then mapped on to the Digital Health Equity framework, which was used to assess the completeness of findings at the different levels and domains of influence (see Table 4). Finally, themes were labelled using data extracts and were organised into two narrative themes, which are presented here.

**Table 4:** Findings aligned with the Digital Health Equity Framework.

|  | **Individual** | **Interpersonal** | **Community** | **Societal** |
| --- | --- | --- | --- | --- |
| **Biological** | Sensory disability (users) |  |  |  |
| **Behavioural** | Accessing care and mental health (users) | Access care autonomously of family (users) |  |  |
| **Physical and built environment** |  | Challenges of home working (providers)<br>Access to private space (users) | Access to clinical space (providers) |  |
| **Digital environment** | Digital self-efficacy (users)<br>Access to technology (users) | Challenges building rapport (both)<br>Safeguarding (providers)<br>Tech bias (providers)<br>Supporting individuals to navigate systems (providers) | Additional work of remote pathways (providers)<br>Drawn-out care (users) | Societal expectations of digital services (both) |
| **Sociocultural environment** | Familiarity with healthcare systems (users)<br>Navigating stigma (users) |  | Remote less prestigious (providers) | Managing stigma of accessing sexual health (users)<br>Managing stigma of being in public (users) |
| **Health system** | Reduced clinic visits (users) |  | Streamlined services (both)<br>Scheduled appointments (both) | Remote prescription (providers) |

### Theme 1: Remote consultations transform the delivery of care

Remote consultations were often part of a wider systemic change in service delivery, either in response to COVID restrictions, or as part of a rationalisation of services and drive to increase efficiency made necessary by reductions in funding and resources. Even though they were not introduced as a stand-alone intervention within services, both professional stakeholders and service users discussed the ways in which they impacted care pathways and delivery of care.

#### “They don’t need to necessarily be here”- Impact on services

Clinicians reported that remote consultations allowed care pathways to be streamlined, prioritising patients who needed in-person services. Coupled with the introduction of OPSS and online advice pathways, remote consultations enabled in-person service provision to be focussed on more complex care needs. For example, asymptomatic service users could often be managed using OPSS and those with simple diagnoses, such as chlamydia, could be managed remotely with medication posted to patients.

Remote consultations could reduce the number of in-person clinic visits for some service users, such as those using PrEP. As one clinician noted, “it’s freeing up appointments that need to be physically seen in comparison to, you know, dropping in to come and get your pills” (HCP12, Healthcare Assistant).

The move to remote consultations was associated with a shift from walk-in services to scheduled appointments. Service managers described how this helped them to better structure services based on capacity.

> *“In the old days we used to have 40 people sitting in a waiting room you know they’d all pile in at half past one when the doors opened and then they would sit there until they were seen… I think if feels more controlled” (HCP07, Nurse/Health Advisor)*

However, another clinician reflected that, although booked appointments allowed services more ability to manage flows of patients, they also introduced inefficiency when patients did not attend their slot.

A senior clinician described how the development of standards supporting remote consultations has re-enabled outreach services run by non-clinical teams to provide treatment to underserved populations in community settings.

> *The outreach team will do the assessment on the clinical system then they’ll phone the prescriber up… if the clinician says, “Yes I’m happy.” They’ll then give the drugs to the person… COVID was definitely helpful for that … before that was almost like, no it can’t be done.” (HCP33, Nurse/Health Advisor)*

Although remote care pathways were often framed in terms of increased efficiency, they could also be associated with additional work, for example the organisation of medication to be posted or picked up. This was particularly pronounced in CSA 3, where some clinicians did remote consultations from home.

> *“Let’s say you have somebody who needs contraception posted to them, when you’re working at home, you can’t do that until you come in the next session… otherwise you have to email your colleague and say, oh, do you mind posting this? But I don’t like putting my work onto somebody else” (HCP04, Nurse/Health Advisor)*

In some cases, uptake of remote services, such as an online PrEP clinic, was lower than expected. This prompted reflection on the need to better understand how people want to engage with SRHS and whether remote services were able to meet service users’ expectations.

> *“I think we are not fast enough… if someone submits their form over the weekend, they might be waiting- you know, five days until it’s processed […] that’s just us not meeting the needs of patients to be efficient or as fast as people are used to - you know - Amazon delivery”. (HCP10, Consultant/Medical Doctor)*

#### “If it was in person, it would have been better” - Impact on service user experience

Both service users and professional stakeholders felt that building rapport and trust during consultations was more challenging remotely. This is a crucial factor in potentially sensitive SRH consultations. However, one service user suggested that rapport can be generated in a remote consultation through clinician’s language and tone of voice.

> *“I think if it’s on the phone, you have to be even, almost even more caring. I feel like the tone of people’s voices means a lot when you do it over the phone because you can’t read someone’s facial expressions. I think I remember saying to my sister, like, when I called sexual health, and then I said, it’s for termination, the woman’s tone completely changed… She’s like, ‘Oh, okay, that’s fine.’ It was really so softly, it made me laugh, but it was actually, really nice.” (SU44, Cis woman 25–34 years old)*

Although video consultations provide some visual cues that could overcome the limitations of a telephone call, some service users reported finding them stressful compared with in-person interactions.

> *“I don’t know why but I feel like with a video call I feel more awkward because I’m on their screen. Whereas, in person, I’m more comfortable because we’re both in person with each other” (SU68, Cis woman, 16–20 years old)*

Some service users expressed a preference for being seen using particular modalities, but many were concerned primarily with the delivery of care, “As long as I felt like they are actually listening to what I’m saying, then it wouldn’t matter what kind of communication” (SU51, Female, 25–34 years old)

Service users highlighted that although a triage and booked appointment system reduces the amount of time waiting in a reception area, they found that seeking care through the new pathways was a longer process.

> *“Pre-Covid when I had symptoms, you just went to the clinic and sat and waited and someone saw you and diagnosed it and treated it. I had [symptoms] again recently, and I felt the process was frustrating […] you couldn’t just go down, you had to ring someone, they had to get back to you then they had to book you in, and it just seemed like a long-winded process. I had to wait three or four days” (SU79, Cis man, 35–44 years old)*

#### “I wouldn’t want to be working in a call centre” - Impact on clinicians

In addition to changes in the mode of interaction, remote consultations also affected clinicians’ job satisfaction. Reflecting the perceived lack of prestige of working remotely, one imagined that only providing remote consultations would be akin to ‘working in a call centre’ (HCP10, Consultant/Medical Doctor).

Those working from home described feeling isolated from colleagues, despite support being available remotely.

> *“It was a big change, when COVID came, to work [from home] … you just feel you haven’t got that, your colleagues around you to ask a question or something even though, you know, you can use Teams, or you can send a message. But it’s not the same.” (HCP04, Nurse/Health Advisor)*

The additional emotional engagement demanded by remote consultations, particularly those relating to abortions, was particularly challenging when working from home.

> *“It’s not a great scenario because you know you’re in your home and so you’re bringing that emotion into your home aren’t you.” (HCP06, Healthcare Assistant)*

This additional burden was not always recognised by their colleagues, and providing remote consultations was sometimes perceived to be less important than seeing patients in clinic.

> *“I don’t think that people who do virtual consultations are seen as working as hard as the ones that have done face-to-face that day. It’s sort of seen as the easier option and you know, it’s not necessarily the easiest option at all.” (HCP03, Nurse/Health Advisor).*

#### “We can’t confirm they’re alone” - Impact on confidentiality

Engaging with service users who were outside the privacy of the clinic room sometimes raised issues of confidentiality for clinicians.

> *“When you try and get hold of them, they’re quite often on the bus or you know, wandering around town or you know, at work or those kinds of things.” (HCP29, Consultant/Medical Doctor)*

Service users described wanting to answer healthcare professional’s questions without fear of being overheard or interrupted, particularly those who shared their living space with family members or friends.

> *“It’s very hard to get a window where you know you’re 100 percent going to be on your own with no distractions. And even if they are out, you’ve also got the worry what happens if they come back early.” (SU84, Cis woman, 21–24 years old)*

The sensitive nature of SRH consultations meant that some clinicians felt that video calls were not always appropriate, particularly when service users joined calls from their bedroom or not fully dressed.

> *“I think it’s important for the client to be able to focus. And also, for their safety, for their privacy as well. And their own modesty, even, that I’m not intruding on them. They wouldn’t come to the clinic in their dressing gown.” (HCP40, Counsellor)*

Issues of confidentiality were intensified when they had concerns about potentially vulnerable service users because they could not guarantee that the person they were speaking to was alone or free from coercion. Concerningly, a clinician reflected that this uncertainty gave colleagues a “get-out clause” to justify not asking safeguarding screening questions that they sometimes found awkward or difficult to engage with in remote consultations.

> *“I think sometimes it’s a bit of a get out clause from asking what can be quite difficult questions and it’s, ‘oh well I couldn’t confirm they were alone because they were over the phone,’ you know. Maybe you should ask regardless but it is a bit of a ways out especially if you’re speaking to someone who’s a little bit confrontational or isn’t the friendliest” (HCP03, Nurse)*

### Theme 2: The challenges and benefits of remote consultation are not distributed evenly

As well as the impact of remote consultations on the way care was provided by SRHS, participants also reflected on the ways in which the remote delivery of care impacted different populations.

#### “The walls are really thin” - Unequal access to private/safe space

A lack of a suitable space made it difficult for some service users to participate in remote consultations. This creates equity issues as certain groups are more likely to live in houses of multiple occupancy and therefore have less access to private or safe spaces. For example, students in shared housing may feel anxious about housemates overhearing consultations.

> *“I know most [students] live in shared housing. As somebody who’s lived in different houses, the walls are really thin, so people can hear phone calls.” (SU75, Non-binary, 16–20 years old)*

A potential lack of privacy also has important implications for the safety of service users experiencing intimate partner violence and safeguarding screening carried out during remote consultations (21). Parents with children may struggle to find a private space at home. SU47 (Cis female, 35–44 years old), described her “loud and manic” home with four children and her concerns about “people listening in while I’m on video call.” However, for other parents, remote access allows them to attend consultations without arranging childcare. Similarly, remote access enables young people living at home to engage with services without having to explain their absence.

> *“You know, I go off cycling [to the clinic], people are going to be asking where are you going? I don’t really like to lie, but that would put me in an awkward situation” (SU58, Cis man, 21–24 years old)*

Remote appointments allowed some participants to access care during working hours without leaving the office. However, these benefits could only be fully realised when their home or work environment provided private time and space. This was often impossible for young people attending school.

> *“For under-18s it’s quite often during school time… There’s been a couple of people that I’ve heard them have arguments with teachers [during a phone consultation] because they’ve tried to go into a quiet classroom and a teacher has come in and said, ‘You can’t be in here on the phone.’” (HCP16, Consultant/Medical Doctor)*

#### “I couldn’t go out the house” - Unequal access to public space

The ability to access services privately was beneficial in communities where being seen accessing an in-person SRHS was associated with greater stigma, including some minoritised ethnic and rural communities. Digital services were perceived by professional stakeholders to mitigate such concerns by offering anonymity and an opportunity to engage with those who were reluctant to attend in-person.

> *“If I were to think of any minority, I would probably say that they may feel more comfortable having maybe the initial telephone consultation to say, “Okay how can we help?” Because there, there is that, that mentality to be like I don’t want to come or, I don’t want to be seen in the clinic.” (HCP35, Nurse/Health Advisor)*

Other communities experiencing marginalisation or discrimination, such as trans and gender diverse people, stood to benefit from being able to access services without potentially exposing themselves to harassment or misgendering by going out in public.

> *“I think worrying about how they might be perceived in public, that could be like a barrier [to in-person clinics] that can be quite difficult for some trans people. Especially early in transition.” (SU52, Trans man, 25–34 years old)*

Remote consultations may improve access to services for individuals with physical or mental health conditions that make attending in person challenging. For example, SU42 explained that a remote consultation could have been beneficial to her when she was experiencing an episode of intense social anxiety.

> *“When I was younger, I had significant mental health problems… there was a year where I couldn’t go out the house. At that point it would have been really beneficial for an on-line consultation to be available to at least make that initial contact” (SU42, Cis woman, 25–34 years old)*

This highlights how having the option for making an initial contact remotely may facilitate access for service users who face initial barriers to attending clinic. These examples highlight how in the digital equity framework, the digital environment can work across individual, community and societal levels to facilitate access to services and overcome some barriers to equity.

#### “Maybe there’s a language barrier” - Individual capacity to engage with remote consultations

Recent migrants and individuals with limited English proficiency may have limited familiarity with the NHS and how to navigate SRHS. Equity was sometimes affected by assumptions and decisions made during triaging, for example, that remote consultations were unsuitable for non-native English speakers.

> *“I had a patient yesterday; she was booked in for a face-to-face abortion consultation. I didn’t have a clue why she was face-to-face because they’re not usually done face-to-face. She was of different ethnicity, so I thought maybe there’s a language barrier. I was all prepped with a language line and all the rest of that… and her English language was very good” (HCP07, Nurse/Health Advisor)*

Physical and sensory difficulties can have an impact on how people interact with services. For example, deafness may preclude telephone use or make remote consultations difficult without support. One service user with visual impairment and hearing loss described how assumptions about their abilities limited the accessibility of remote consultation.

> *“People presume[e] if you’re unable to see or have trouble seeing you can make phone calls. Rather than I have trouble seeing <u>and</u> I have trouble with phone calls.” (SU66, Non-binary, 25–34 years old)*

Other participants described how issues, such as poor memory, made remote consultations possible but challenging. This shaped their preferences for how they engaged with services.

> *“My preference is always, always, always for face-to-face consultations. On a phone consultation I feel rushed… because my memory is so bad that I just have to have notes to know what I want to talk about… [In person] If I’m getting good eye contact from the doctor when I’m mentioning something, it cues me to remember something else.” (SU10, Cis man, over 45 years old)*

Thus, remote consultations can enhance equity for users with physical, cognitive, or mental health conditions, but only if implemented with accessible options.

#### “They come in to speak to us at the desk ” – Navigating digital barriers

While digital services may increase uptake, they require digital literacy, self-efficacy, and confidence. Services relying heavily on digital pathways risk excluding those with lower digital literacy. In two services studied, appointments had to be booked via a single web or telephone pathway. Although no formal alternatives existed, front-line staff often supported service users to navigate systems, which was, although not an official part of their role, time-consuming.

> *“They come in to speak to us at the desk because they don’t, they can’t access these things. We have people come in; they want to make an appointment. “How do I do this on my phone?” (HCP24, Healthcare Assistant)*

Access to and the cost of digital technology affect service users’ ability to engage in remote appointments. Interview data showed that participation depends on having smartphones or tablets and reliable internet or Wi-Fi. In CSA3, which served a wide geographical area, the provision of remote consultations improved accessibility, but poor internet and mobile connectivity remained a barrier. Additionally, the sensitive nature of SRH can make it difficult for patients to share devices with others.

Individuals who have limited access to mobile technology were not able to engage with digitised processes, such as receiving test results via text.

> *“We do have like an older cohort of 70, 80-year-olds and they definitely prefer to come to clinic. And then when we say, you’re going to have results by text, they say well I don’t have any way to get a text, that doesn’t work for me.” (HCP19, Consultant/Medical Doctor)*

Although older people were often perceived by other participants to be a group who may lack digital literacy, older participants we interviewed felt able to navigate online systems. Nevertheless, digital literacy cannot be assumed, and some individuals may need additional support to navigate systems regardless of their age.

> *“You’ve got people that are dyslexic You’ve got people[who’ve] got autism, you’ve got people with learning difficulties, and this goes, it’s not stereotyping to any age. This is right across the board” (HCP24, Healthcare Assistant)*

Conversely, young people were frequently perceived to be most comfortable with digital systems and believed to prefer remote consultations. However, the personal nature of SRHS meant that young service users prioritised face-to-face consultation. One participant mentioned that they did not think they would be able to get the same level of detail from a remote appointment.

> *“I would say with sexual matters being a very personal thing, people – I’d say myself as well – prefer to speak about that to someone as opposed to a screen or through their phone.” (SU33, Cis man, 21–24 years old)*

Other young people found service websites old fashioned or difficult to navigate, so preferred walk-in services.

> *“When it comes to sexual health, I prefer to just walk into the clinic because I found that booking a time on the website for the clinic, on the NHS website, is quite complicated, the website is a bit outdated.” (SU31, Non-binary, 16–20 years old)*

Across professional stakeholder and service user interviews, young people were presumed to have greater fluency with technology. However, they sometimes lacked the resources to be able to access services digitally.

> *“There are different versions of remote. So even down to you send a text message to somebody but they’re on a pay-as-you-go phone, they’ll get charged for the text message. If they’re a youngster and I know this because we’ve been told by them, they, they can’t afford to get the message” (HCP27, Manager)*

Many services had already developed alternative pathways and outreach for marginalised service users in partnership with support networks, community venues, and pharmacies. This investment in outreach and alternative provision acknowledges the existing inequities in in-person SRHS provision. The data in this section overlay these provisions and suggest that the introduction of remote consultation has potentially eased access for some populations less able, or less willing, to access in-person services but at the expense of access for those experiencing other marginalisations.

## DISCUSSION

This analysis has shown how remote consultations have transformed the delivery of care. While helping to improve efficiency, remote pathways have also introduced additional clinical work, such as managing the distribution of medicines, and appear to have negatively impacted job satisfaction for some clinicians. Although acceptable, remote consultations were recognised to impact the quality of interaction during consultations, affecting rapport between service users and clinicians. For some service users, remote pathways were longer and more challenging to navigate than walk-in models. Importantly, taking place outside of the ‘safe space’ of the clinical room, clinicians were unable to assure service user privacy, with implications for confidentiality and safeguarding.

Applying the digital health equity framework, our findings illustrate that the benefits and challenges of remote consultations are unevenly distributed, potentially introducing new, or exacerbating existing health inequalities. Although remote consultations can facilitate access for those unable or unwilling to attend clinics in person, they require users to have privacy, technology, and the capacity to engage in telephone or video consultations. Applying the framework shifts focus from individual capacities and emphasises the multiple levels of influence on the digital environment navigated by service users and providers. This highlights structural and digital barriers that may disproportionately affect certain groups.

Language barriers were sometimes perceived by professional stakeholders to preclude remote consultations. Although such judgements may be based on previous experiences with consultations, generalising about people’s capacity risks undermining access and equity. Our research adds to the growing body of evidence on the use of remote consultations in healthcare and extends it beyond general practice and into SRH. The findings align with studies from other settings, which have shown that remote consultations can be adequate for managing common problems (22) and may also be more efficient than in-person consultations (23). However, challenges remain, including digital inequalities, reduced quality of care, difficulties in building rapport, and increased clinician stress (22–24). Building on this broader evidence, our findings highlight that the acceptability of remote consultations may vary between populations. Although young people were often perceived to be a key population for remote consultation, many young people we spoke to expressed a preference for attending consultations in person. Evidence from other studies of this age group is mixed. A study of online postal self-sampling for chlamydia testing in England found lowest rates of uptake among 15–19 year-olds, but highest overall engagement among those 20–24 years old. A cross-sectional study of 905 young people in Australia identified that 75% preferred OPSS to in-person STI testing, citing the greater privacy and convenience of on-line testing. However, those who preferred in-clinic testing felt this approach was more trustworthy (25). The accessibility and trustworthiness of services was also highlighted as a key consideration by young people in a focus group study of 16-18-year-olds’ perceptions of digital sexual health services (26).

Our findings suggest that while remote modalities may play a key role in providing convenient access to STI screening and information, in-person services remain a key place to deliver trustworthy care in a safe clinical space. Our data highlights a lack of safe, private space as a key concern for some groups accessing remote consultation. Although not included in our sample, these concerns are likely to affect people on low incomes, refugees and asylum seekers, people experiencing housing instability, and those in temporary accommodation who could face similar challenges. Older adults were also a population highlighted by participants as preferring in-person consultations, which resonates with the findings of a UK discrete-choice experiment in which participants over the age of 45 years, who have often been neglected in sexual health research, prioritised face-to-face consultations (27). A further study of the experiences of adults over 45 accessing SRHS in England highlighted the intersection of disability and age and noted the physical inaccessibility of services was a significant barrier for older disabled people (28). While remote consultations can help to provide some services without the need to physically access service premises, our data highlight gaps in provision of remote pathways for people with intersecting physical and sensory disabilities. Our analysis suggests that the intersection of disability with other forms of marginalisation, for example the high rates of neurodivergence and musculoskeletal comorbidities among the trans community (29), makes remote access to services potentially more important for some groups. The unpredictability of long-term conditions highlights the need for flexible consultation formats, allowing switches between in-person and remote.

The importance of considering intersecting barriers to remote engagement are also highlighted in our data and are supported by a study of older adults from disadvantaged backgrounds with limited English accessing primary care video consultations (12). In addition, the loss of cues shifts the burden of being understood on to patients and a recent study suggests that marginalised patients may struggle to express symptoms over the phone, risking misdiagnosis and exclusion (30). These studies emphasise how remote consultations can exacerbate existing inequalities and reduce patient agency by introducing barriers to engagement. Services in our study had engaged with the needs of underserved communities and engaged in non-digital outreach to provide services in community settings, with evidence in one service that the adoption of remote prescription and distribution of medication had enhanced the outreach offering. However, our findings showed that frontline service staff sometimes acted unofficially as the interface between services users and redefined digital pathways, highlighting the need for greater accessibility and support for underserved communities (31). These findings demonstrate the intersection of age, disability, and socioeconomic position, emphasising that digital pathways may either facilitate or hinder access depending on personal, social, and structural factors, a core consideration of the digital health equity framework.

For some, digital services may align well with proactive and autonomous health-seeking behaviours, offering convenience and discretion. For others, the availability of familiar face-to-face provision may limit engagement with new service offerings. In the context of the NHS 10-year health plan, there is proposed expansion of digital technologies across health services, with an emphasis on digital-first service interactions wherever clinically appropriate. This includes remote triage, consultations, and ongoing monitoring through virtual platforms (1). While such innovations have the potential to improve access, our findings underline the importance of recognising that digital pathways are not equally accessible or appropriate for all. This reinforces the framework of digital health equity and the interactions between technology and social and structural determinants.

The findings of this study also highlight the need for continued research into the evolving landscape of service delivery. The COVID-19 pandemic was a key driver in the rapid expansion of remote consultation, yet the configuration of services continues to shift. For example, one CSA in this study modified its service delivery model during the study period, limiting remote consultations to specific circumstances. This illustrates the fluidity of service provision, and the need for ongoing evaluation of how remote consultations are adopted and adapted. Future research should explore the equity implications of these changes, particularly in relation to access for marginalised and underserved populations. From an implementation perspective, ensuring that demand for services is not assumed and communities are meaningfully engaged in the design and delivery of services to support ease of use and acceptability, applying frameworks developed for remote consultation ethically, is critical (32).

This study benefits from the inclusion of both service users and professional stakeholders from three diverse CSAs to gain insight into the successes and challenges of remote consultations from a variety of perspectives, including the experiences of members of marginalised communities. However, by primarily recruiting through clinical services, we reached few service users who had never attended in-person services and those who had never accessed SRHS, particularly those from underserved communities. Those with only remote or no experiences of care may have different perspectives.

## CONCLUSION

By engaging with service users, potential service users and professional stakeholders from three diverse case study areas, this study demonstrates the multifaceted role that remote consultations play in shaping equitable access to SRHS in England and Wales, its impact on service delivery and clinicians, and the factors involved in delivering that care. Applying the framework of digital health equity, our findings highlight that the benefits and burdens of remote care are unevenly distributed, for instance, increased convenience for some coexisted with exclusion for those lacking private space or reliable connectivity, reflecting structural, social, and individual factors that influence access and engagement. Remote consultations may inadvertently widen health inequalities due to varying access to privacy, technology, and digital literacy. Continued research on the implementation of and evaluation of the impact of remote consultation use in SRHS is needed to understand how evolving models of remote care affect equity, ensuring that services are actively designed and responsive to community needs and do not inadvertently widen health inequalities.

## Data Availability

All data produced in the present study are available upon reasonable request to the authors

